# Awareness and Acceptance of Deep Margin Elevation amongst Dental Practitioners- A cross-sectional multicentric study

**DOI:** 10.64898/2026.01.16.26344250

**Authors:** Jitendra Jethwani, Gayathri Sundari, Lovely M Annamma, Esam Tashkandi, Caroline K Carrico

**Affiliations:** Virginia Commonwealth University, School of Dentistry, Richmond, Virginia, USA; Department of Restorative Dentistry, Saveetha Dental College, Chennai, India; Department of Prosthodontics, College of Dentistry, King Saud University, Riyadh, Saudi Arabia

**Keywords:** Deep margin elevation, restorative dentistry, cervical margin relocation, coronal margin relocation, proximal box elevation

## Abstract

**Background:** Deep margin elevation (DME) is a restorative technique that facilitates the placement of restorations in cases of subgingival margins. Although clinically reported, very few data are available on dental practitioners’ awareness and use of DME.

**Objectives:** To evaluate awareness and clinical acceptance toward deep margin elevation (DME) use in subgingival restorative cases among dental practitioners worldwide.

**Methodology:** A cross-sectional questionnaire-based study was conducted among practicing dentists at various dental educational institutions, private dental practices, and a combination of academic and private dental practices across multiple centers globally. The self-administered questionnaire consisted of 20 closed-ended questions to evaluate awareness and clinical acceptance. The data were entered into and analyzed using a Chi-square test and descriptive statistics in the Statistical Package for the Social Sciences (SPSS) software.

**Results:** Out of 450 invited participants, 349 general dental practitioners completed the survey (77.6%). The purely educational institutions’ response rate was 79 (23%), the strictly private dental practice response rate was 134 (39%), and the combined academic and private practice response rate was 131 (38%). Sixty-six percent of respondents agreed that predictable adhesive bonding to cervical/root dentin can be achieved in restorations with deep margins. Although a majority of respondents had heard of DME (77%), the majority reported a preference for surgical crown lengthening (75%) when favorable conditions were present.

**Conclusion:** The study highlights moderate DME awareness among the study participants. The findings of this study revealed that the number of dentists who use the technique to restore large subgingival defects in posterior teeth with proximal caries is very small. Thus, it is recommended that dental practitioners introduce this technique in their dental clinics as an alternative to surgical crown lengthening. Although years of experience and a dentist’s rank may influence clinical decisions, an in-depth factorial analysis with a larger sample size is necessary.

## Introduction

Advancements in restorative dentistry focused on achieving functional longevity and biological compatibility among the various restorative materials used for different procedures. The most complex restorative procedures are deep carious lesions, restorations that repeatedly fail and restorations with deep subgingival margins, especially in posterior teeth. The reasons for difficulty in subgingival margin placement include isolation, impression accuracy, and biologic width encroachment, which can alter gingival tissue and periodontal health (1). Reliable, durable adhesion to dentin with newer dentin adhesives revolutionized adhesive restorations. Still, they were not without problems when the margins of the lesion extended beyond the CEJ or sub-gingivally. The other reason was that proper marginal adaptation of subgingival restorations was difficult, which led to marginal leakage, staining, and sensitivity after restoration placement (2). Dietschi and Spreafico proposed the initial concept of deep margin elevation in 1998 (3), but the idea was later reported and popularized as a solution for clinical crown lengthening by Magne et al. (4). The synonyms for deep margin elevation (DME) are cervical margin relocation, proximal box elevation (PBE), open sandwich technique and coronal margin relocation (CMR) (5,6,7). DME involves placing a composite resin layer at the base of class II subgingival restorations in posterior teeth to elevate it to the gingival or supragingival level (7). This technique’s advantages include good accessibility, better isolation, enhanced bonding, preservation of periodontal health, minimal invasiveness, and protection of the pulp (7). The DME is technique-sensitive and, if not meticulously performed or if inappropriate restorative materials are used, can lead to microleakage and periodontal issues (8). Before the DME technique, the popular methods used to increase the crown-root ratio for prosthodontic restorative procedures were surgical crown-lengthening (SCL) (9) and orthodontic extrusion of the clinical crowns (10). In SCL, marginal bone reduction was often necessary to provide adequate distance from the bone crest to the cervical margin of the restoration (11). The disadvantage of this technique is that it is a surgically invasive procedure, and, as attachment loss occurs, the furcation and root concavities of the involved tooth may become exposed (4). The surgical crown-lengthening procedure (SCL) led to tissue loss, root exposure, opening of the proximal area (leading to black holes), and poor aesthetics. Orthodontic extrusion is time-consuming, can cause root resorption, and can lead to oral hygiene issues when wearing the orthodontic appliance (12). Due to the complications and invasive procedures associated with SCL and orthodontic extrusion, the DME shifted the paradigm for treating deep cavities (13). This study aims to determine the awareness and acceptance of DME among dental practitioners, restorative/general dentists and periodontists. This study hypothesized that although a surgical crown-lengthening procedure (SCL) is valuable, its indications should decrease over time, given that DME, despite being a very demanding procedure, appears to be well tolerated by the surrounding periodontium, clinically and histologically.

## Methodology

This research used a self-administered questionnaire validated by experts in restorative dentistry. The study used a cross-sectional survey to assess awareness and clinical acceptance of deep-margin elevation among practicing dental professionals from the Middle East, the USA, and Europe. A cross-sectional survey was selected to evaluate the global trend of clinical acceptance. The study was approved by VCU IRB, and the ethical approval number was HM20023469.

A questionnaire survey was circulated to purely private practice, academic institutions, and a combination of educational and dental practice settings. The inclusion criteria were licensed dental practitioners in the three regions. An informed consent form for participation was attached to the questionnaire. Participants were excluded if they were unwilling to participate in the survey or had difficulty understanding English. The target sample size was calculated to be 350 based on the participant awareness level (65%) as per local data (14), a 95% confidence interval, and a 5% margin of error (nQuery v9.3.1, Statistical Solutions, Boston, MA). The survey was distributed by convenience sampling through a snowball sampling method. The validated self-administered questionnaire collected the data. The questionnaire had three main sections: demographics, awareness of DME, and clinical acceptance. The demographics included age, gender, professional qualification, and experience in the type of dental practice setting. The questions based on awareness and clinical acceptance were the second section. A total of 20 closed-end questions evaluated awareness and clinical acceptance of the DME technique. The questionnaire was sent as a pilot survey to a panel of experts (15 n) to assess content validity and make modifications. After the validation and reliability checks, the questionnaire was distributed in hard copy, via email, and via WhatsApp, depending on participants’ accessibility. Electronic survey responses were collected and managed with Research Electronic Data Capture (REDCap) hosted at Virginia Commonwealth University. REDCap is a secure, web-based software platform designed to support data capture for research studies (15). The questionnaire was sent to dental institutions, private practices, and academic and dental practice settings, and to known colleagues in the Middle East, Europe, and the USA. Participation was voluntary, and confidentiality was maintained during data collection. Two reminders were sent via email or phone. The data was qualitatively and quantitatively evaluated. All collected data were checked for completeness. None of the questions were mandatory, and the missing data ranged from 0 to 12 respondents. Responses were summarized as counts and percentages based on the number of respondents to each question. Associations between awareness and clinical acceptance questions and respondent characteristics, including practice setting (academic, private practice, combination) and practice location (Middle East/Asia vs Other), were examined using chi-squared tests. SAS EG v.8.3 (SAS Institute, Cary, NC, USA) was used for all analyses. A significance level of 0.05 was considered to be statistically significant.

## Results

A total of 349 general dentists were included in the analysis. More than half of respondents reported practicing in the Middle East (n=184, 53%), followed by the United States (n=54, 15%) and Asia (n=48, 14%), with the remaining respondents from Europe and other regions. There was a nearly equal split among respondents: 134 (39%) in purely private practice, 131 (38%) in a combination of academic and private practice, and 79 (23%) in a purely academic practice setting. Some respondents identified additional training, including prosthodontics (n=50, 14%), AEGD (n=42, 12%), or operative dentistry (n=43, 12%). A complete summary of respondent characteristics is provided in Table 1.

**Table 1.**
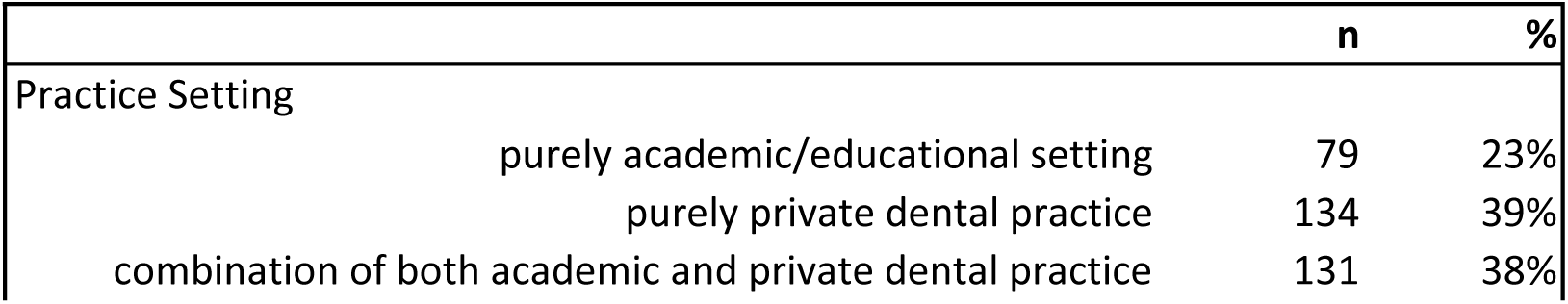
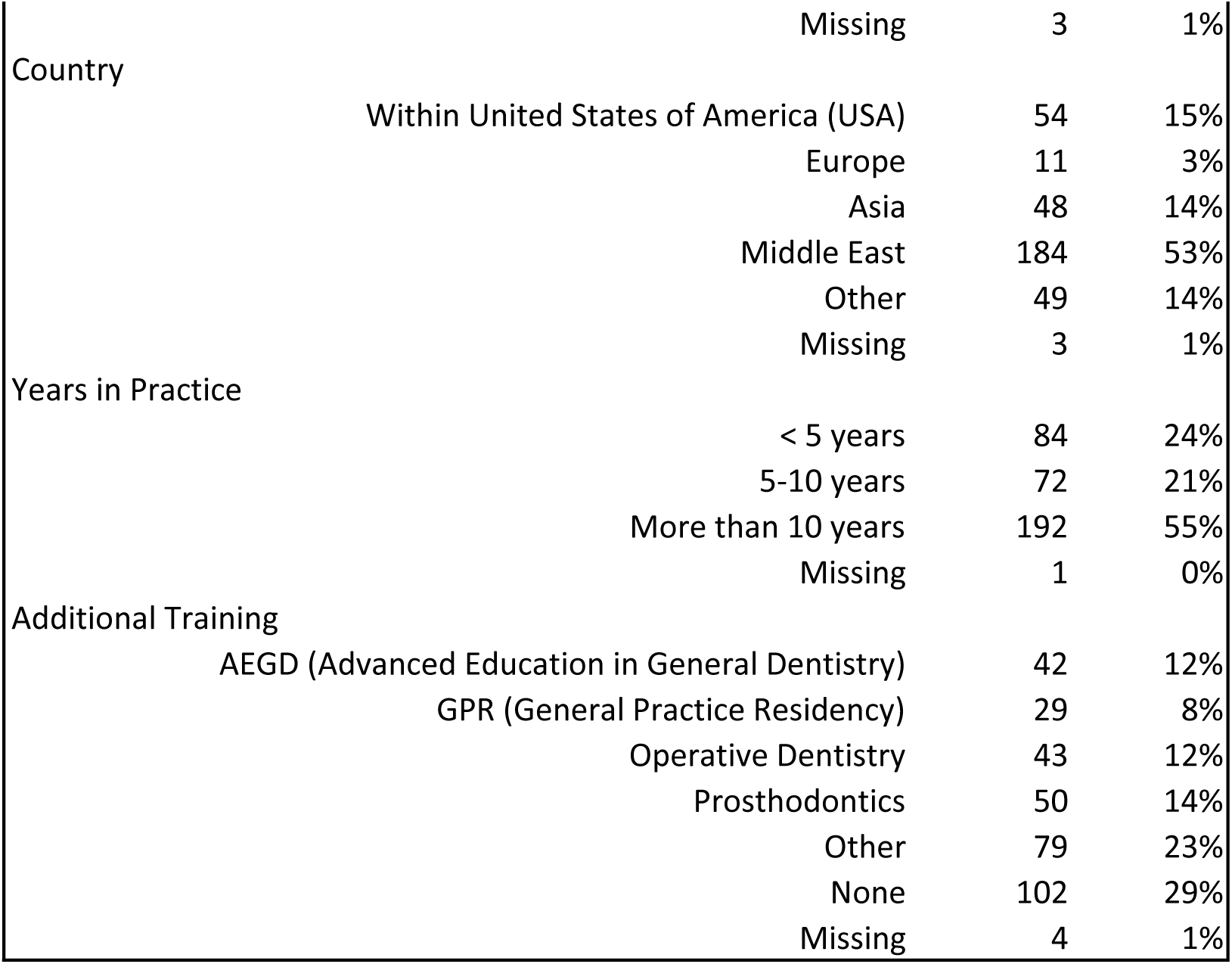
Respondent Characteristics.

A summary of the responses to statements regarding DME is provided in Table 2. More than half of respondents indicated a belief that predictable adhesive bonding to cervical/root dentin in restorations with deep margins can be achieved (n=228, 66%). More than 75% of respondents had heard of the deep marginal elevation technique (including by other terminology: cervical margin elevation, proximal box elevation, cervical margin relocation, open sandwich technique) (n=268, 77%). Eighty-five per cent of respondents agreed that the gingival location of the restoration margins determines the success of a restoration by influencing marginal adaptation (n=293, 85%). Bitewing (n=225, 65%) and intraoral periapical radiographs (n=89, 26%) were selected more frequently than Orthopantomographic radiographs (n=32, 9%) to view the subgingival marginal adaptation of a restoration. A 2mm standard biological width was most frequently reported by respondents as the minimum required to support a restoration, and nearly all respondents (n=331, 96%) agreed that it is best clinical practice to evaluate existing biological width before choosing DME in cavities with subgingival margins. A majority of respondents agreed that DME will improve clinical longevity of a tooth (n=288, 83%) and improve the success of final restoration (n=287, 84%), and only 49% agreed that DME can cause restoration failure due to poor cervical margin adaptation. Overall, 79% preferred DME with good isolation and definitive restoration over orthodontic extrusion or surgical crown lengthening (n=271, 79%). In the presence of an ideal crown: root ratio (1:1.5), only 54% (n=188) would prefer orthodontic extrusion, but surgical crown lengthening was indicated by 75% (n=260) as preferred over DME if favorable conditions were present. Over 80% agreed that presenting complications of orthodontic extrusion, such as root resorption and tooth fracture, should be included. Surgical crown lengthening complications included attachment loss and furcation exposure. (84%, 83%, respectively). For teeth requiring endodontic treatment, there was no clear consensus on the timing of DME: 42% preferred before, 37% after, and 21% did both procedures in a single visit.

**Table 2.**
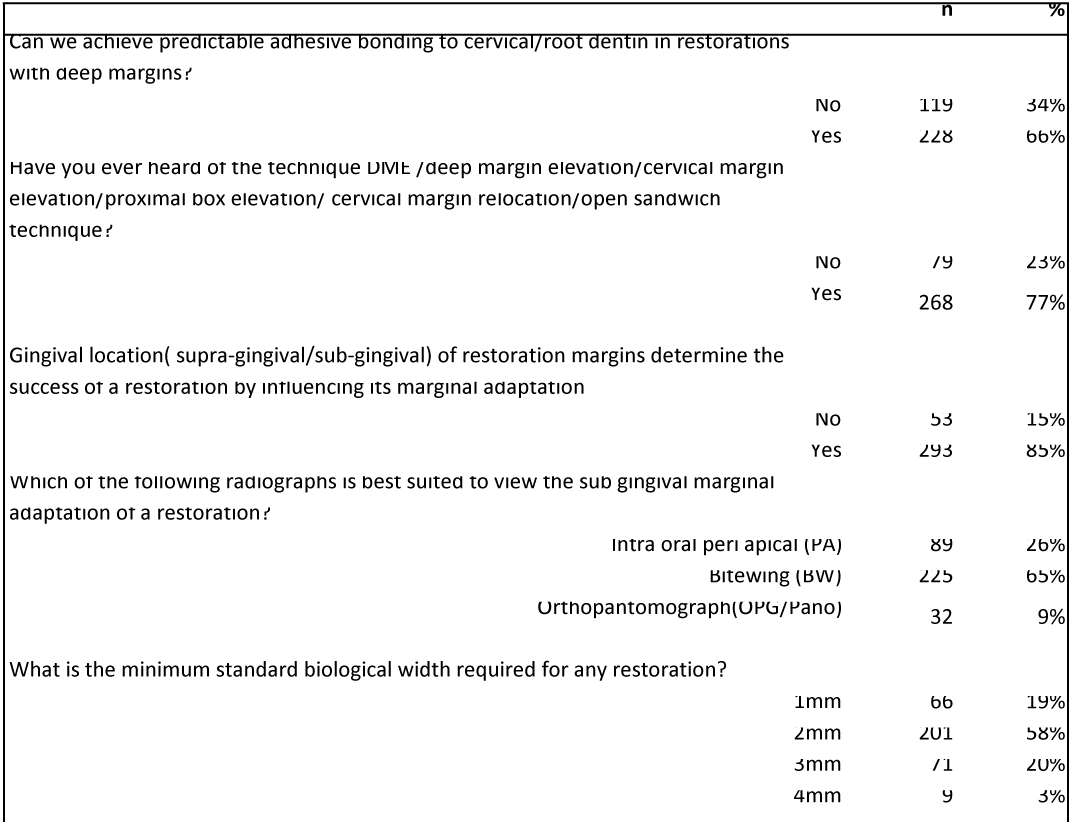
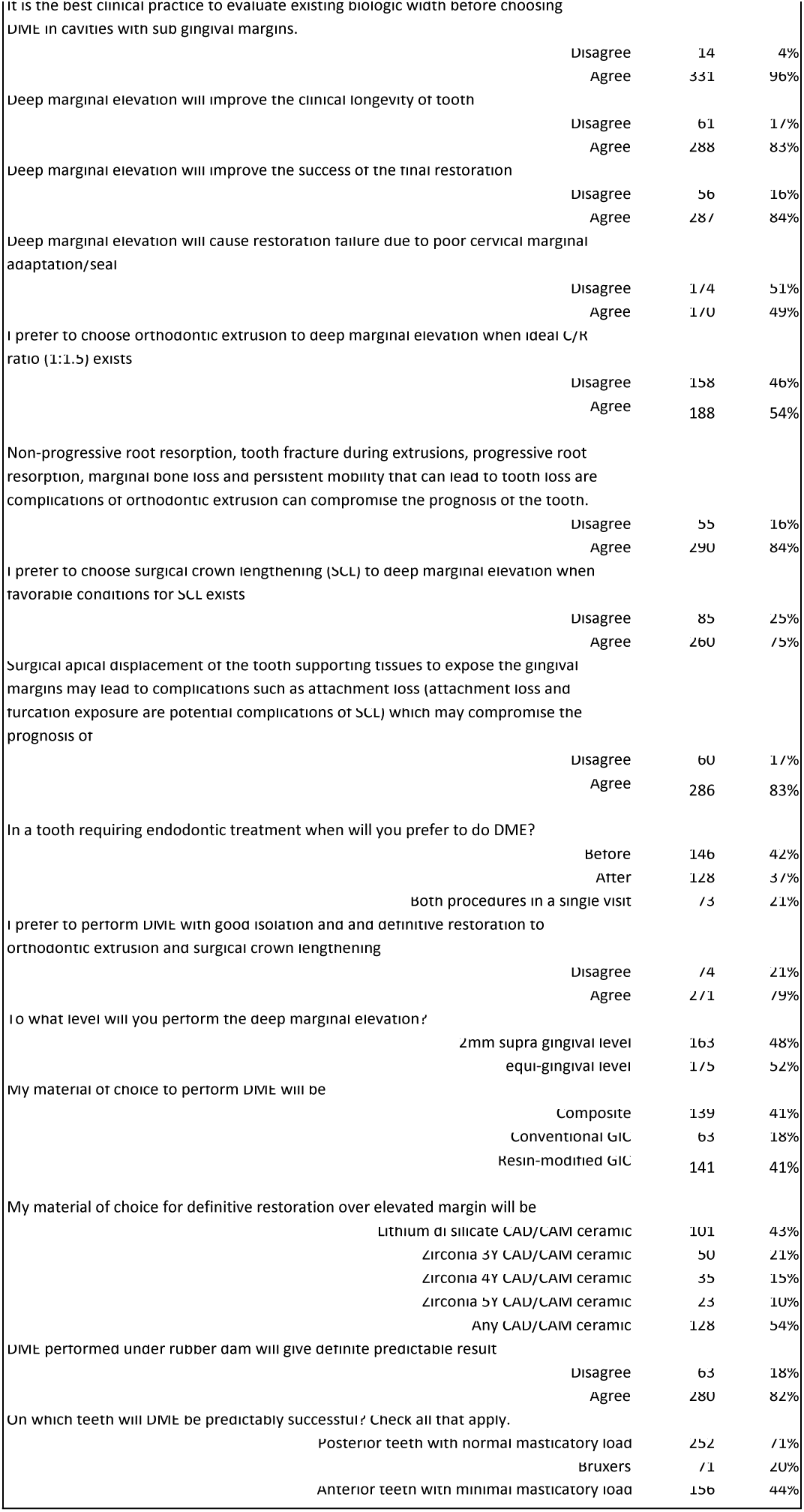
Summary of Practice Preferences.

|  | n | % |
| --- | --- | --- |
| Can we achieve predictable adhesive bonding to cervical/root dentin in restorations with deep margins? |  |  |
| No | 119 | 34% |
| Yes | 228 | 66% |
| Have you ever heard of the technique DME: /deep margin elevation/cervical margin elevation/proximal box elevation/ cervical margin relocation/open sandwich technique? |  |  |
| No | 79 | 23% |
| Yes | 268 | 77% |
| Gingival location( supra-gingival/sub-gingival) of restoration margins determine the success of a restoration by influencing its marginal adaptation |  |  |
| No | 53 | 15% |
| Yes | 293 | 85% |
| Which of the following radiographs is best suited to view the sub gingival marginal adaptation of a restoration? |  |  |
| Intra oral peri apical (PA) | 89 | 26% |
| Bitewing (BW) | 225 | 65% |
| Orthopantomograph(OPG/Pano) | 32 | 9% |
| What is the minimum standard biological width required for any restoration? |  |  |
| 1mm | 66 | 19% |
| 2mm | 201 | 58% |
| 3mm | 71 | 20% |
| 4mm | 9 | 3% |

|  |  |  |  |
| --- | --- | --- | --- |
| It is the best clinical practice to evaluate existing biologic width before choosing DME in cavities with sub gingival margins. |  |  |  |
|  | Disagree | 14 | 4% |
|  | Agree | 331 | 96% |
| Deep marginal elevation will improve the clinical longevity of tooth |  |  |  |
|  | Disagree | 61 | 17% |
|  | Agree | 288 | 83% |
| Deep marginal elevation will improve the success of the final restoration |  |  |  |
|  | Disagree | 56 | 16% |
|  | Agree | 287 | 84% |
| Deep marginal elevation will cause restoration failure due to poor cervical marginal adaptation/seal |  |  |  |
|  | Disagree | 174 | 51% |
|  | Agree | 170 | 49% |
| I prefer to choose orthodontic extrusion to deep marginal elevation when ideal C/R ratio (1:1.5) exists |  |  |  |
|  | Disagree | 158 | 46% |
|  | Agree | 188 | 54% |
| Non-progressive root resorption, tooth fracture during extrusions, progressive root resorption, marginal bone loss and persistent mobility that can lead to tooth loss are complications of orthodontic extrusion can compromise the prognosis of the tooth. |  |  |  |
|  | Disagree | 55 | 16% |
|  | Agree | 290 | 84% |
| I prefer to choose surgical crown lengthening (SCL) to deep marginal elevation when favorable conditions for SCL exists |  |  |  |
|  | Disagree | 85 | 25% |
|  | Agree | 260 | 75% |
| Surgical apical displacement of the tooth supporting tissues to expose the gingival margins may lead to complications such as attachment loss (attachment loss and furcation exposure are potential complications of SCL) which may compromise the prognosis of |  |  |  |
|  | Disagree | 60 | 17% |
|  | Agree | 286 | 83% |
| In a tooth requiring endodontic treatment when will you prefer to do DME? |  |  |  |
|  | Before | 146 | 42% |
|  | After | 128 | 37% |
|  | Both procedures in a single visit | 73 | 21% |
| I prefer to perform DME with good isolation and and definitive restoration to orthodontic extrusion and surgical crown lengthening |  |  |  |
|  | Disagree | 74 | 21% |
|  | Agree | 271 | 79% |
| To what level will you perform the deep marginal elevation? |  |  |  |
|  | 2mm supra gingival level | 163 | 48% |
|  | equi-gingival level | 175 | 52% |
| My material of choice to perform DME will be |  |  |  |
|  | Composite | 139 | 41% |
|  | Conventional GIC | 63 | 18% |
|  | Resin-modified GIC | 141 | 41% |
| My material of choice for definitive restoration over elevated margin will be |  |  |  |
|  | Lithium di silicate CAD/CAM ceramic | 101 | 43% |
|  | Zirconia 3Y CAD/CAM ceramic | 50 | 21% |
|  | Zirconia 4Y CAD/CAM ceramic | 35 | 15% |
|  | Zirconia 5Y CAD/CAM ceramic | 23 | 10% |
|  | Any CAD/CAM ceramic | 128 | 54% |
| DME performed under rubber dam will give definite predictable result |  |  |  |
|  | Disagree | 63 | 18% |
|  | Agree | 280 | 82% |
| On which teeth will DME be predictably successful? Check all that apply. |  |  |  |
|  | Posterior teeth with normal masticatory load | 252 | 71% |
|  | Bruxers | 71 | 20% |
|  | Anterior teeth with minimal masticatory load | 156 | 44% |

When performing DME, the preferred level of elevation was split between 2mm supra-gingival (n=163, 48%) and equigingival (n=175, 52%). The preferred material was predominantly composite (n=139, 41%) or resin-modified glass ionomers (n=141, 41%), with fewer preferring conventional glass ionomers (n=63, 18%). For the definitive restoration over an elevated margin, Lithium disilicate CAD/CAM ceramic was most commonly indicated (n=101, 43%).

A majority of respondents felt that posterior teeth with normal masticatory load would be predictably successful with DME (n=252, 71%), with fewer agreeing about anterior teeth with minimal masticatory load (n=156, 44%) and in patients with a history of bruxism (n=71, 20%).

Many beliefs and practices with deep marginal elevation were significantly associated with the provider’s practice setting (Table 3). Specifically, academic respondents showed a higher level of agreement that predictable adhesive bonding to cervical/root dentin in restorations with deep margins can be achieved (77% vs 65% and 58%, p-value=0.0182). Although the majority of respondents indicated 2mm as the minimum standard biological width required for restorations (51% to 66%), academic respondents were more likely to indicate 1mm as the minimum (n=24, 31% vs. 18-20% for private practice, combination) and those in private practice or combination of private and academic were more likely to indicate 3mm (19%-26% vs 13%) (p-value=0.0028). Significantly more academic providers also agreed that DME will cause restoration failure due to poor cervical marginal adaptation or seal (63% vs. 45-46%, p-value=0.0282) and that orthodontic extrusion is preferable when the ideal crown: root ratio (1:1.5) is present (75% vs 44-53%, p-value<0.0001). Additionally, academic providers were more likely to prefer performing DME with good isolation and definitive restoration rather than orthodontic extrusion or surgical crown lengthening (95% vs 73-79%, p-value=0.0170). The preferred level of DME was 2mm supragingival for 68% of those in academic settings compared to 40-44% for those in some or entirely private practice who had a higher rate of agreement with equigingival level (55-60% vs 32%) (p-value=0.0006). Academic providers also had a greater preference for Lithium di silicate CAD/CAM ceramic materials (44%) while those in private practice or a combination were more likely to indicate any CAD/CAM ceramic (42-43%) (p-value=0.0045). Nearly all academic respondents agreed that DME performed under a rubber dam will give definite, predictable results, but fewer of those who work in private practice or a combination (97% vs 76-80%, p-value=0.0003). Finally, respondents in an academic setting were less likely to indicate anterior teeth with minimal masticatory load as teeth where DME would be predictably successful (31% vs 47-50%, p-value=0.0223).

**Table 3.**
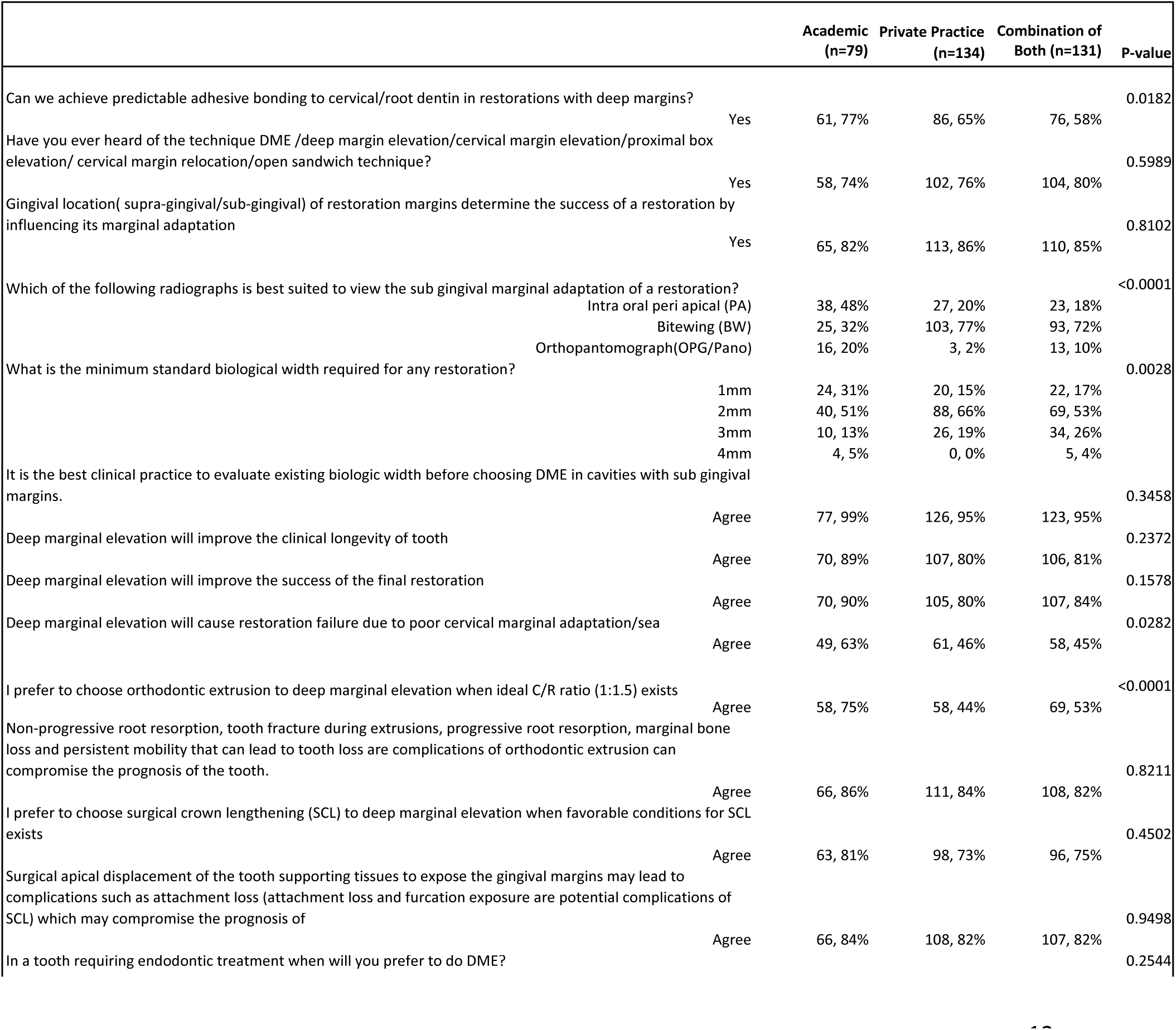
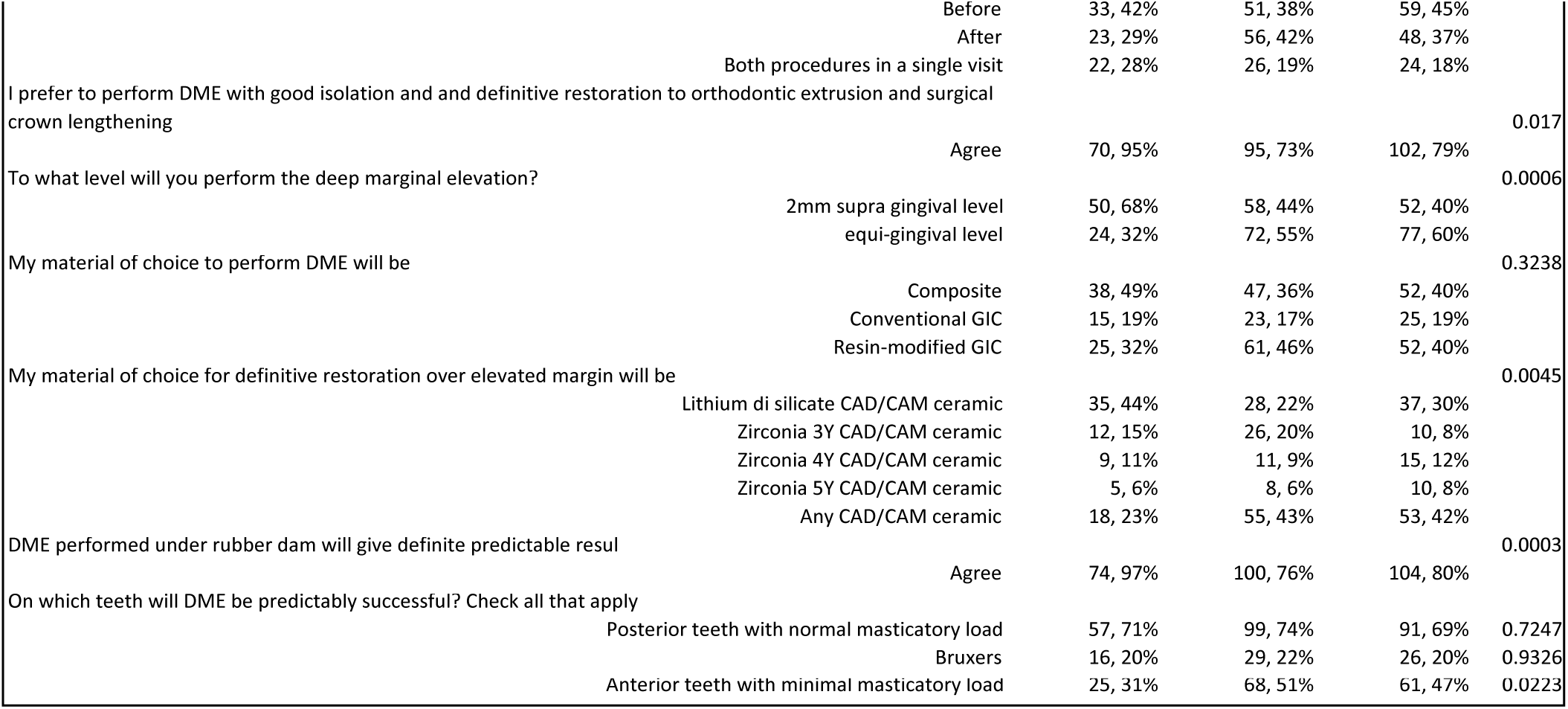
Provider Type Associations.

Respondents were also categorized into two groups based on their geographical region, with 232 from the Middle East and Asia (67%) and the rest grouped as “Other”, which included the US and Europe (n=114, 33%). There were also self-reported beliefs and practices based on these geographical distinctions (Table 4). Although both groups responded favorably overall, those from the Middle East or Asia were significantly more likely to agree that DME will improve the clinical longevity of a tooth (86% vs 76%, p-value=0.0289). Middle Eastern and Asian respondents were also slightly more likely to prefer orthodontic extrusion over DME when the ideal crown: root ratio exists. Middle Eastern and Asian respondents were also more likely to agree that DME performed under a rubber dam will give definite, predictable results (87% vs 71%, p-value=0.0005). Finally, providers from Europe and the United States were more likely to feel anterior teeth with minimal masticatory load were predictably successful with DME (54% vs 41%, p-value=0.0048). There were marginal differences in perceptions regarding DME in bruxers (p-value=0.0613) and in preference for DME over orthodontic extrusion and surgical crown lengthening (p-value=0.0675).

**Table 4.** Practice Region Associations.

|  |  | Middle<br>East/Asia<br>(n=232) | Other (n=114) | P-value |
| --- | --- | --- | --- | --- |
| Can we achieve predictable adhesive bonding to cervical/root dentin in restorations with deep margins? |  |  |  | 0.4336 |
|  | Yes | 155, 67% | 71, 63% |  |
| Have you ever heard of the technique DME /deep margin elevation/cervical margin elevation/proximal box elevation/<br>cervical margin relocation/open sandwich technique? |  |  |  | 0.2924 |
|  | Yes | 174, 76% | 92, 81% |  |
| Gingival location( supra-gingival/sub-gingival) of restoration margins determine the success of a restoration by<br>influencing its marginal adaptation |  |  |  | 0.3017 |
|  | Yes | 199, 87% | 93, 82% |  |
| Which of the following radiographs is best suited to view the sub gingival marginal adaptation of a restoration? |  |  |  | 0.4989 |
|  | Intra oral peri apical (PA) | 62, 27% | 25, 22% |  |
|  | Bitewing (BW) | 146, 63% | 79, 70% |  |
|  | Orthopantomograph(OPG/Pano) | 22, 10% | 9, 8% |  |
| What is the minimum standard biological width required for any restoration? |  |  |  | 0.8842 |
|  | 1mm | 43, 19% | 22, 19% |  |
|  | 2mm | 135, 58% | 65, 58% |  |
|  | 3mm | 48, 21% | 22, 19% |  |
|  | 4mm | 5, 2% | 4, 4% |  |
| It is the best clinical practice to evaluate existing biologic width before choosing DME in cavities with sub gingival<br>margins. |  |  |  | 0.6545 |
|  | Agree | 222, 97% | 107, 96% |  |
| Deep marginal elevation will improve the clinical longevity of tooth |  |  |  | 0.0289 |
|  | Agree | 199, 86% | 87, 76% |  |
| Deep marginal elevation will improve the success of the final restoration |  |  |  | 0.9298 |
|  | Agree | 191, 83% | 93, 84% |  |
| Deep marginal elevation will cause restoration failure due to poor cervical marginal adaptation/seal |  |  |  | 0.3745 |
|  | Agree | 116, 51% | 51, 46% |  |
| I prefer to choose orthodontic extrusion to deep marginal elevation when ideal C/R ratio (1:1.5) exists |  |  |  | 0.0392 |
|  | Agree | 133, 58% | 52, 46% |  |
| Non-progressive root resorption, tooth fracture during extrusions, progressive root resorption, marginal bone loss<br>and persistent mobility that can lead to tooth loss are complications of orthodontic extrusion can compromise the<br>prognosis of the tooth. |  |  |  | 0.8351 |
|  | Agree | 192, 84% | 95, 83% |  |
| I prefer to choose surgical crown lengthening (SCL) to deep marginal elevation when favorable conditions for SCL<br>exists |  |  |  | 0.2669 |
|  | Agree | 177, 77% | 80, 71% |  |
| Surgical apical displacement of the tooth supporting tissues to expose the gingival margins may lead to<br>complications such as attachment loss (attachment loss and furcation exposure are potential complications of<br>SCL) which may compromise the prognosis of |  |  |  | 0.8576 |
|  | Agree | 190, 82% | 93, 83% |  |
| In a tooth requiring endodontic treatment when will you prefer to do DME? |  |  |  | 0.2313 |
|  | Before | 104, 45% | 40, 35% |  |
|  | After | 80, 35% | 47, 42% |  |
|  | Both procedures in a single visit | 47, 20% | 26, 23% |  |
| I prefer to perform DME with good isolation and and definitive restoration to orthodontic extrusion and surgical<br>crown lengthening |  |  |  | 0.0675 |
|  | Agree | 186, 81% | 82, 73% |  |
| To what level will you perform the deep marginal elevation? |  |  |  | 0.7171 |
|  | 2mm supra gingival level | 111, 49% | 51, 47% |  |
|  | equi-gingival level | 116, 50% | 58, 51% |  |
| My material of choice to perform DME will be |  |  |  | 0.1850 |
|  | Composite | 90, 39% | 47, 43% |  |
|  | Conventional GIC | 49, 21% | 14, 13% |  |
|  | Resin-modified GIC | 93, 40% | 48, 44% |  |
| My material of choice for definitive restoration over elevated margin will be |  |  |  | 0.9413 |
|  | Lithium di silicate CAD/CAM ceramic | 66, 29% | 33, 31% |  |
|  | Zirconia 3Y CAD/CAM ceramic | 36, 16% | 14, 13% |  |
|  | Zirconia 4Y CAD/CAM ceramic | 23, 10% | 11, 10% |  |
|  | Zirconia 5Y CAD/CAM ceramic | 17, 7% | 6, 6% |  |
|  | Any CAD/CAM ceramic | 87, 38% | 41, 39% |  |
| DME performed under rubber dam will give definite predictable result |  |  |  | 0.0005 |
|  | Agree | 198, 87% | 80, 71% |  |
| On which teeth will DME be predictably successful? Check all that apply. |  |  |  |  |
|  | Posterior teeth with normal masticatory load | 161, 69% | 88, 77% | 0.1291 |
|  | Bruxers | 41, 18% | 30, 26% | 0.0613 |
|  | Anterior teeth with minimal masticatory load | 94, 41% | 62, 54% | 0.0048 |

## Discussion

The present study on clinicians’ perceptions, knowledge, and clinical decision-making regarding DME provided valuable insights into current trends, preferences, and clinical approaches. Most studies on awareness, knowledge, and application of DME were conducted as single-institution or single-country studies. The uniqueness of this study lies in its multicentric design. In our study, the categories were also classified into academic and private settings, and into private practitioners and academicians, which provides valuable insight into variation in clinical decision-making patterns by practice type. A high level of DME technique awareness was observed in 80% of participants, which aligns with similar findings reported by (14) and (16). The adhesive bonding to cervical root/dentin was a concern in our study amongst private and in combination practitioners, though the academicians believed it to be achievable. Nonetheless, studies showed that recent adhesive systems can achieve durable bonds in deep cervical areas with optimal isolation. (17). For radiographic evaluation, bitewing radiographs were preferred due to their accurate visualization of proximal margins and adaptation of class II restorations. (18) Another key factor, the biological width required, varied from 2 to 3mm amongst the practitioners for minimum standards for restorations. The ideal is 2.5 to 3mm, with an average of 2mm. (19) Most respondents 80% above agree that DME improves clinical longevity and success of restoration, which is a finding similar to other studies. (4) & (7) The clinical alternatives to DME, such as orthodontic extrusion and surgical crown lengthening, carry the risk of root resorption and loss of attachment, which was a common finding in other studies. Among the two alternative procedures, the majority preferred surgical crown lengthening over orthodontic extrusion. (13) (20) The preferred time for doing DME is before the endodontic procedure, as recommended by studies (7, 21)

The preferred level of elevation, as per our study, was 2mm supragingival or equigingival. This finding is similar to other studies for better isolation and bonding while maintaining periodontal health. (22) Among the choices of restorative material for DME resin-modified glass ionomers (RMGIC) and composite was the most common. RMGIC releases fluoride and exhibits good chemical adhesion, whereas composite resin provides superior marginal integrity and aesthetics when proper isolation methods are used. (23,24). The clinical implications observed indicate that DME is widely accepted as a minimally invasive technique for managing subgingival margins with no adverse effects, as in orthodontic extrusion or surgical crown lengthening. Differences in choices between the private practitioners and academicians largely relate to variations in experience and risk control rather than disagreement regarding the validity of DME.

### Limitations of the study

The study is self-reported and may introduce bias or overestimate clinical proficiency. A cross-sectional study captures data at a single point in time and does not account for cumulative experience. Thirdly, the data were collected across multiple centers, and the majority of respondents were from the Middle East; hence, the findings cannot be generalized to other regions. The specialist category was not a major one, leading to an imbalance in clinical perspectives, particularly regarding biologic width and periodontal considerations.

## Conclusion

Within the limitations of the study, it can be concluded that, though most clinicians widely recognize and accept DME as the most viable minimally invasive option, the variability in confidence regarding bonding predictability and margin adaptation suggests a need for hands-on training to improve clinical consistency.

Most clinicians believe that DME enhances restoration success and longevity, provided that the biologic width is respected and proper isolation methods are achieved.

For future research, more prospective clinical trials comparing DME, surgical crown lengthening, and orthodontic extrusion are required to assess periodontal stability, marginal adaptation, and the long-term survival of restorations.

In vivo studies evaluating histologic and microbiologic effects of DME on safe margins with multiple composite resins and resin-modified glass ionomers will confirm safety, along with evidence-based clinical guidelines.

## Data Availability

All relevant data are within the manuscript and its Supporting Information files.

## Acknowledgement

The authors are thankful to the Saudi Society of Periodontology for their technical support

## References

1. Veneziani M. Adhesive restorations in the posterior area with subgingival cervical margins: new classification and differentiated treatment approach. Eur J Esthet Dent. 2010;5(1):50–76. PMID: 20305873

2. Mahrous AI, Eltiti HA, Ahmed IM, Alagha EI. Effect of different gingival margin restorations of class II cavities on microleakage: an in-vitro study. Electron Physician. 2015 Nov 20;7(7):1435–40. doi: 10.19082/1435. PMID: 26767095; PMCID: PMC4700887.

3. Dietschi D, Spreafico R. Current clinical concepts for adhesive cementation of tooth-colored posterior restorations. Pract Periodontics Aesthet Dent. 1998 Jan-Feb;10(1):47–54; quiz 56. PMID: 9582662

4. Magne P, Spreafico RC. Deep margin elevation: a paradigm shift. Am J Esthet Dent. 2012 Jun 1;2(2):86–96.

5. Grubbs TD, Vargas M, Kolker J, Teixeira EC. Efficacy of direct restorative materials in proximal box elevation on the margin quality and fracture resistance of molars restored with CAD/CAM onlays. Oper Dent. 2020 Jan-Feb;45(1):52–61. doi: 10.2341/18-098-L. Epub 2019 May 14. PMID: 31084532.

6. Butt A. Cervical margin relocation and indirect restorations: case report and literature review. Dent Update. 2021 Feb;48(2):93–100. doi: 10.12968/denu.2021.48.2.93.

7. Aldakheel M, Aldosary K, Alnafissah S, Alaamer R, Alqahtani A, Almuhtab N. Deep margin elevation: current concepts and clinical considerations: a review. Medicina (Kaunas). 2022 Oct 18;58(10):1482. doi: 10.3390/medicina58101482. PMID: 36295642; PMCID: PMC9610387

8. Miles DA. A taxonomy of research gaps: identifying and defining the seven research gaps. J Res Methods Strateg. 2017;1:1–15.

9. Al-Sowygh ZH. Does surgical crown lengthening procedure produce stable clinical outcomes for restorative treatment? A meta-analysis. J Prosthodont. 2019;28(1):103–109. doi: 10.1111/jopr.12909.

10. Cordaro M, Staderini E, Torsello F, Grande NM, Turchi M, Cordaro M. Orthodontic extrusion vs. surgical extrusion to rehabilitate severely damaged teeth: a literature review. Int J Environ Res Public Health. 2021 Sep 10;18(18):9530. doi: 10.3390/ijerph18189530. PMID: 34574454; PMCID: PMC8469087

11. Padbury Jr A, Eber R, Wang HL. Interactions between the gingiva and the margin of restorations. J Clin Periodontol. 2003;30(5):379–385. doi: 10.1034/j.1600-051X.2003.01277.x.

12. Bach N, Baylard JF, Voyer R. Orthodontic extrusion: periodontal considerations and applications. J Can Dent Assoc. 2004 Dec;70(11):775–80. PMID: 15588553.

13. Sarfati A, Tirlet G. Deep margin elevation versus crown lengthening: biologic width revisited. Int J Esthet Dent. 2018;13(3):334–356. PMID: 30073217

14. Binalrimal SR, Banjar WM, Alyousef SH, Alawad MI, Alawad GI. Assessment of knowledge, attitude, and practice regarding Deep Margin Elevation (DME) among dental practitioners in Riyadh, Saudi Arabia. J Family Med Prim Care. 2021;10(5):1931–1937. doi:10.4103/jfmpc.jfmpc_1707_20

15. Harris PA, Taylor R, Thielke R, Payne J, Gonzalez N, Conde JG. Research electronic data capture(REDCap)-A metadata-driven methodology and workflow process for providing translational research informatics support. J Biomed Inform. 2009;42(2):377–381.doi:10.1016/j.jbi.2008.08.010

16. Padaru, Mythri, et al. “Awareness and Practice of Deep Margin Elevation among Dental Practitioners in India: A Cross-Sectional Survey.” Pesquisa brasileira em odontopediatria e clínica integrada 25 (2025):

17. Kitahara, S., Shimizu, S., Takagaki, T., Inokoshi, M., Abdou, A., Burrow, M. F., & Nikaido, T. (2024). Dentin Bonding Durability of Four Different Recently Introduced Self-Etch Adhesives. Materials, 17(17), 4296. 10.3390/ma17174296

18. Signori C, Laske M, Mendes FM, Huysmans MDNJM, Cenci MS, Opdam NJM. Decision-making of general practitioners on interventions at restorations based on bitewing radiographs. J Dent. 2018 Sep;76:109–116. doi: 10.1016/j.jdent.2018.07.003. Epub 2018 Jul 9. PMID: 30004002.

19. Ingber JS, Rose LF, Coslet JG. The “biologic width”—a concept in periodontics and restorative dentistry. Alpha Omegan. 1977;70:62–5

20. Karageorgiou, A., Fostiropoulou, M., Antoniadou, M., & Pappa, E. (2025). Deep Margin Elevation: Current Evidence and a Critical Approach to Clinical Protocols—A Narrative Review. Adhesives, 1(3), 10. 10.3390/adhesives1030010

21. Da Silva D., Ceballos L., Fuentes M. Influence of the Adhesive Strategy in the Sealing Ability of Resin Composite Inlays After Deep Margin Elevation. J. Clin. Exp. Dent. 2021;8:e886–e893. doi: 10.4317/jced.58689.

22. Hausdörfer T, Lechte C, Kanzow P, Rödig T, Wiegand A. Periodontal health in teeth treated with deep-margin-elevation and CAD/CAM partial lithium disilicate restorations-a prospective controlled trial. Clin Oral Investig. 2024 Nov 30;28(12):670. doi: 10.1007/s00784-024-06053-y. PMID: 39613879; PMCID: PMC11606998.

23. Brenes-Alvarado A, Cury JA. Fluoride Release from Glass Ionomer Cement and Resin-modified Glass Ionomer Cement Materials under Conditions Mimicking the Caries Process. Oper Dent. 2021 Jul 1;46(4):457–466. doi: 10.2341/19-296-L. PMID: 34478544.

24. Arbildo-Vega HI, Cruzado-Oliva FH, Coronel-Zubiate FT, Luján-Valencia SA, Meza-Málaga JM, Aguirre-Ipenza R, Echevarria-Goche A, Luján-Urviola E, Castillo-Cornock TB, Serquen-Olano K. Comparison of clinical performance of glass ionomer cement vs. composite resin in restorations of non-carious cervical lesions: A systematic review and meta-analysis. J Clin Exp Dent. 2025 Aug 1;17(8):e995–e1005. doi: 10.4317/jced.62997. PMID: 40950519; PMCID: PMC12424605.

